# High-risk exposure without personal protective equipment and infection with SARS-CoV-2 in healthcare workers: results of the CoV-CONTACT prospective cohort

**DOI:** 10.1101/2020.09.17.20194860

**Authors:** Sarah Tubiana, Charles Burdet, Nadhira Houhou, Michael Thy, Pauline Manchon, François Blanquart, Charlotte Charpentier, Jérémie Guedj, Loubna Alavoine, Sylvie Behillil, Anne Leclercq, Jean-Christophe Lucet, Yazdan Yazdanpanah, Mikaël Attia, Caroline Demeret, Thierry Rose, Julia Anna Bielicki, Patricia Bruijning-Verhagen, Herman Goossens, Diane Descamps, Sylvie van der Werf, Bruno Lina, Xavier Duval

**Affiliations:** AP-HP, Hôpital Bichat, Centre d’Investigation Clinique, Inserm CIC 1425, F-75018 Paris, France; Université de Paris, IAME, INSERM, F-75018 Paris, France; AP-HP, Hôpital Bichat, Centre de Ressources Biologiques, F-75018 Paris, France; AP-HP, Hôpital Bichat, Département d’Epidémiologie, Biostatistique et Recherche, F-75018 Paris, France; AP-HP, Hôpital Bichat, Laboratoire de Virologie, F-75018 Paris, France; Center for Interdisciplinary Research in Biology (CIRB), Collège de France, CNRS, INSERM, PSL Research University, Paris, France; Molecular Genetics of RNA Viruses, Department of Virology, CNRS UMR3569, Université de Paris, Institut Pasteur, Paris, France; National Reference Center for Respiratory Viruses, Institut Pasteur, Paris, France; AP-HP, Beaujon Hospital, Direction des soins, F-92118 Clichy, France; AP-HP, Hôpital Bichat, Equipe de Prévention du Risque Infectieux, F-75018 Paris, France; AP-HP, Hôpital Bichat, Service de Maladies Infectieuses et tropicales, F-75018 Paris, France; Physique des fonctions biologiques, CNRS UMR3738, Institut Pasteur, Paris, France; Biologie cellulaire des lymphocytes, INSERM – U1221, Department of Immunology, Institut Pasteur, Paris, France; Paediatric Infectious Diseases Research Group, Institute for Infection and Immunity, St George’s University of London, London SW17 ORE, UK; Paediatric Pharmacology and Paediatric Infectious Diseases, University of Basel Children’s 5Hospital, Basel, Switzerland; Julius Center for Health Sciences and Primary Care, University Medical Center Utrecht, Utrecht, The Netherlands; Laboratory of Medical Microbiology, Vaccine and Infectious Disease Institute, Faculty of Medicine and Health Science, University of Antwerp, Antwerp, Belgium; CIRI, Centre International de Recherche en Infectiologie, (Team VirPath), Univ Lyon, Inserm, Ullll, Université Claude Bernard Lyon 1, CNRS, UMR5308, ENS de Lyon, F-69007, Lyon, France; Laboratoire de Virologie, Centre National de Référence des Virus des infections respiratoires (dont la grippe), Institut des Agents Infectieux, Groupement Hospitalier Nord, Hospices Civils de Lyon, 69004, Lyon, France

**Keywords:** SARS-CoV-2, healthcare workers, high-risk exposure, personal protective equipment, transmission

## Abstract

**Objective:** We aimed to estimate the risk of infection in Healthcare workers (HCWs) following a high-risk exposure without personal protective equipment (PPE).

**Methods:** We conducted a prospective cohort in HCWs who had a high-risk exposure to SARS-CoV-2-infected subject without PPE. Daily symptoms were self-reported for 30 days, nasopharyngeal swabs for SARS-CoV-2 RT-PCR were performed at inclusion and at days 3, 5, 7 and 12, SARS-CoV-2 serology was assessed at inclusion and at day 30. Confirmed infection was defined by positive RT-PCR or seroconversion, and possible infection by one general and one specific symptom for two consecutive days.

**Results:** Between February 5^th^ and May 30^th^, 2020, 154 HCWs were enrolled within 14 days following one high-risk exposure to either a hospital patient (70/154; 46.1%) and/or a colleague (95/154; 62.5%). At day 30, 25.0% had a confirmed infection (37/148; 95%CI, 18.4%; 32.9%), and 43.9% (65/148; 95%CI, 35.9%; 52.3%) had a confirmed or possible infection. Factors independently associated with confirmed or possible SARS-CoV-2 infection were being a pharmacist or administrative assistant rather than being from medical staff (adjusted OR (aOR)=3.8, CI95%=1.3;11.2, p=0.01), and exposure to a SARS-CoV-2-infected patient rather than exposure to a SARS-CoV-2-infected colleague (aOR=2.6, CI95%=1.2;5.9, p=0.02). Among the 26 HCWs with a SARS-CoV-2-positive nasopharyngeal swab, 7 (26.9%) had no symptom at the time of the RT-PCR positivity.

**Conclusions:** The proportion of HCWs with confirmed or possible SARS-CoV-2 infection was high. There were less occurrences of high-risk exposure with patients than with colleagues, but those were associated with an increased risk of infection.

## Introduction

Since December 2019, severe acute respiratory syndrome coronavirus 2 (SARS-CoV-2) which causes COVID-19, rapidly spread around the globe [1, 2]. The difficulty to control its rapid propagation is related to many factors, including the fact that infectiousness can precede the symptoms onset, thereby complicating the identification and isolation of infected individuals before they can transmit the virus [3,4].

Healthcare workers (HCWs) can be infected following a contact with a patient, but also after interactions with colleagues, or in the community [5-12]. The use of personal protective equipment (PPE) was rapidly implemented in, departments hosting suspected or identified SARS-CoV-2-infected subjects. However, the atypical presentation of the infection favors high risk contacts between HCW and unidentified patients in other departments [3], and the large circulation of the virus increases the risk of infection during interactions with colleagues, during which the use of PPE or social distancing may be less strictly followed [13].

We conducted a prospective cohort study to estimate the risk of infection in HCWs following high-risk exposure in the hospital, and to evaluate the virological, immunological and clinical outcomes following exposure.

## Methods

### Study design and participants

The CoV-CONTACT study is an ongoing prospective multicenter cohort study including HCWs with exposure to an "index" SARS-CoV-2-infected person (either a patient or a colleague) whose infection was virologically proven by a nasopharyngeal RT-PCR and whose exposure was considered at high-risk of SARS-CoV-2 transmission. HCWs were included in the study in the 14 days following the last identified high-risk exposure. The present analysis focuses on "contact" subjects enrolled at the >1000 bed Bichat Claude Bernard University Hospital (Paris, France) [14] between March, 3^rd^ 2020 and April, 27^th^ 2020.

### Ethics and regulatory issues

The study was approved by the French National Data Protection Commission (approval #920102), and the French Ethics committee (CPP-Ile-de-France-6, #2020-A00280-39) and was registered on the Clinicaltrial.gov registry (NCT04259892). All subjects provided written informed consent.

### Definition of high-risk exposure

Exposure was considered to be at high-risk of SARS-CoV-2 transmission if it occurred i) face-to-face, within one meter and without protective surgical or FFP2/N95 mask, and ii) during a discussion or while the index had an episode of coughing or sneezing, and iii) in the 72 hours prior to, or following the virological diagnosis, or during the symptomatic period of the index.

### Data collection

Collected characteristics of the index included age, date of the diagnostic nasopharyngeal RT-PCR and the SARS-CoV-2 viral load [15].

The collected characteristics of the contacts included medical history, weight, height, current medications, and smoking status. The date of the last high-risk exposure, D0, with the index were also recorded, as well as the cumulative exposure duration.

Contacts were followed up for 30 days following D0. Nasopharyngeal swabs were performed at inclusion, and then at D3, D5, D7 and D12. As inclusion could occur up to 14 days after D0, a maximum of five nasopharyngeal swabs could be collected. Blood samples were drawn at inclusion and D30±7d for SARS-CoV-2 serology.

A set of general symptoms (fever >38°C, fatigue, myalgia, headache) and specific symptoms (cough, breathing difficulties, sore throat, nasal congestion, anosmia, diarrhea) was recorded daily from D0 to D30 using self-administered questionnaires. Results of any additional nasopharyngeal swab were collected, as well as the occurrence of hospitalization, or the existence of household contacts hospitalized for a SARS-CoV-2 infection between D0 and D30.

### Virology

The SARS-CoV-2 RT-PCR was performed blinded to contact characteristics and reported symptoms (see Supplementary appendix).

In contacts with clinical signs suggestive of COVID-19 but a negative SARS-CoV-2 RT-PCR and a negative SARS-CoV-2 serology at D30, a multiplex RT-PCR (QIAstat-Dx Respiratory Panel; Qiagen, Germany) was retrospectively performed on available aliquots to detect other respiratory pathogens (see Supplementary appendix).

### Serology

SARS-CoV-2 serology was performed blinded to contacts’ characteristics and reported symptoms. We used two methods targeting different SARS-CoV-2 antigens: LuLISA N, an in-house Luciferase-Linked Immunosorbent assay designed to detect IgG targeted toward SARS-CoV-2 N antigen (unpublished results) and EuroIMMUN, a commercial immunoassay used for the detection of IgG targeted toward the SARS-CoV-2 recombinant Spike protein subunit (S1) [16]. A serum was considered as positive for SARS-CoV-2 antibodies when the signal exceeded the threshold set at 13,402 relative light units per second (RLU/s) for LuLISA or a 1.1 ratio for EuroIMMUN.

For each method, we defined SARS-CoV-2 seroconversion as the apparition of a positive SARS-CoV-2 serology at the D30 visit, or as an at least two-fold increase of the LuLISA signal or EuroIMMUN ratio between inclusion and D30, in the case of a positive serology at inclusion.

### Definition of SARS-CoV-2 infection

Three definitions of SARS-CoV-2 infection were used: (i) "clinically-suspected infection", when the contact reported at least one general symptom and one specific symptom during two consecutive days during the 30-day follow-up; (ii) "virologically-proven infection", if the contact had at least one SARS-CoV-2-positive nasopharyngeal swab during the 30-day follow-up; (iii) "immunologically-proven infection" if the contact exhibited a SARS-CoV-2 seroconversion in any of the two methods.

SARS-CoV-2 infection was considered as confirmed if it was virologically or immunologically-proven, and considered as possible in case of clinically-suspected infection only.

The primary endpoint was confirmed or possible SARS-CoV-2 infection, thereafter referred to as SARS-CoV-2 infection.

### Statistical methods

Categorical variables are expressed as counts (percentage) and continuous variables are expressed as median (IQR). We first estimated the prevalence of SARS-CoV-2 infection among HCWs, with its 95% confidence intervals computed using the binomial distribution. For the primary endpoint, in case of missing data for one of the components, the subject was considered as infected if one of the available components of the endpoint fulfilled the definition of infection, and considered missing otherwise.

We searched for risk factors associated with SARS-CoV-2 infection among HCWs. Variables achieving a p-value <0.20 in univariate logistic regression analysis were entered into a multivariate logistic regression analysis. Using a backward selection method, we obtained a final model in which all risk factors had a p-value <0.05.. A sensitivity analysis was performed after exclusion of the subpopulation who only met the definition of a possible SARS-CoV-2 infection.

We then studied the kinetics of the SARS-CoV-2 infection in participants with virologically-proven and clinically-suspected infection. We analyzed the SARS-CoV-2 viral load as a function of time from symptom onset using a quadratic regression model..

Analyses were performed with R v3.5 (R Foundation for Statistical Computing, Vienna, Austria). All tests were two-sided with a type-1 error fixed to 0.05.

## Results

### Contacts characteristics and type of exposure

Overall, 154 HCWs exposed to 44 COVID-19 index subjects were included. The median age of these contacts was 35 years (IQR 29.0; 46.8), 35/154 were male (22.7 %). High-risk exposure occurred prior to the widespread use of masks in the hospital (on March, 18^th^) in 88/154 contacts (57%). In contrast, the exposure to colleagues increased from 31.8% (28/88) before March, 18^th^ to 88.2% (60/68) after March, 18^th^ (2 contacts with combined exposures). Overall 28/154 contacts (18.2%) had a high-risk exposure with more than one index subject.

The median duration of the high-risk exposure period was two days (IQR 0; 3.8), and contacts were enrolled at a median time of 6.5 days (IQR 4; 8) after D0. Table 1 presents the characteristics of the included contacts; 51/154 (33.1%) were medical doctors, midwifes, or residents; 77/154 (50.0%) were nurses, nursing assistant, physiotherapists or hospital students.

Contacts were exposed to patients (70/152, 46.1%) or colleagues (95/152, 62.5 %) of whom 13/152 (8.6%) were exposed to both patients and colleagues.

While the exposure to patients represented 68.2% (60/88) of high-risk exposure before March 18^th^, this number dropped to only 11.8% (8/68) after March 18^th^. Most contacts had a cumulated exposure of more than 30 minutes (102/151, 67.5%). In the 95 contacts exposed to an index colleague, exposure was related to face-to-face discussion (89/95, 93.7%), meetings (26/95, 27.4%), lunch sharing (20/95, 21.1%), and other (10/95,10.5%).

### Virological, immunological and clinical outcomes

Overall, 26/154 contact subjects (16.9%, 95%CI [11.3%;23.8%]) had at least one SARS-CoV-2-positive nasopharyngeal swab (see details in supplementary appendix). When positive, the median nasopharyngeal SARS-CoV-2 viral load was 8.7 logio copies/ml (IQR 6.5; 9.4).

Overall, 147 of the 154 contacts had both inclusion and D30 sera samples. At inclusion, 15/147 (10%) contacts had a positive serology by one of the two methods. At D30, 31/147 (21.1%, 95%CI [14.8%; 28.6%]) contacts exhibited a seroconversion. Results obtained by the two serological methods are presented in Table S1.

Based on self-administered questionnaires, 61/151 (40.4%, 95%CI [32.5%; 48.7%]) contacts met the definition of a clinical infection (see details in supplementary appendix). The median duration of symptoms was 5.5 days (IQR 3; 9.2).

The Figure 1 presents the combination of virological, immunological and clinical outcomes; 28 contacts fulfilled only the clinical definition of infection. In these subjects, the prevalence of symptoms dropped at day 10, whereas it persisted elevated until day 30 in those with confirmed infection (Figure 2).

**Figure 1.**
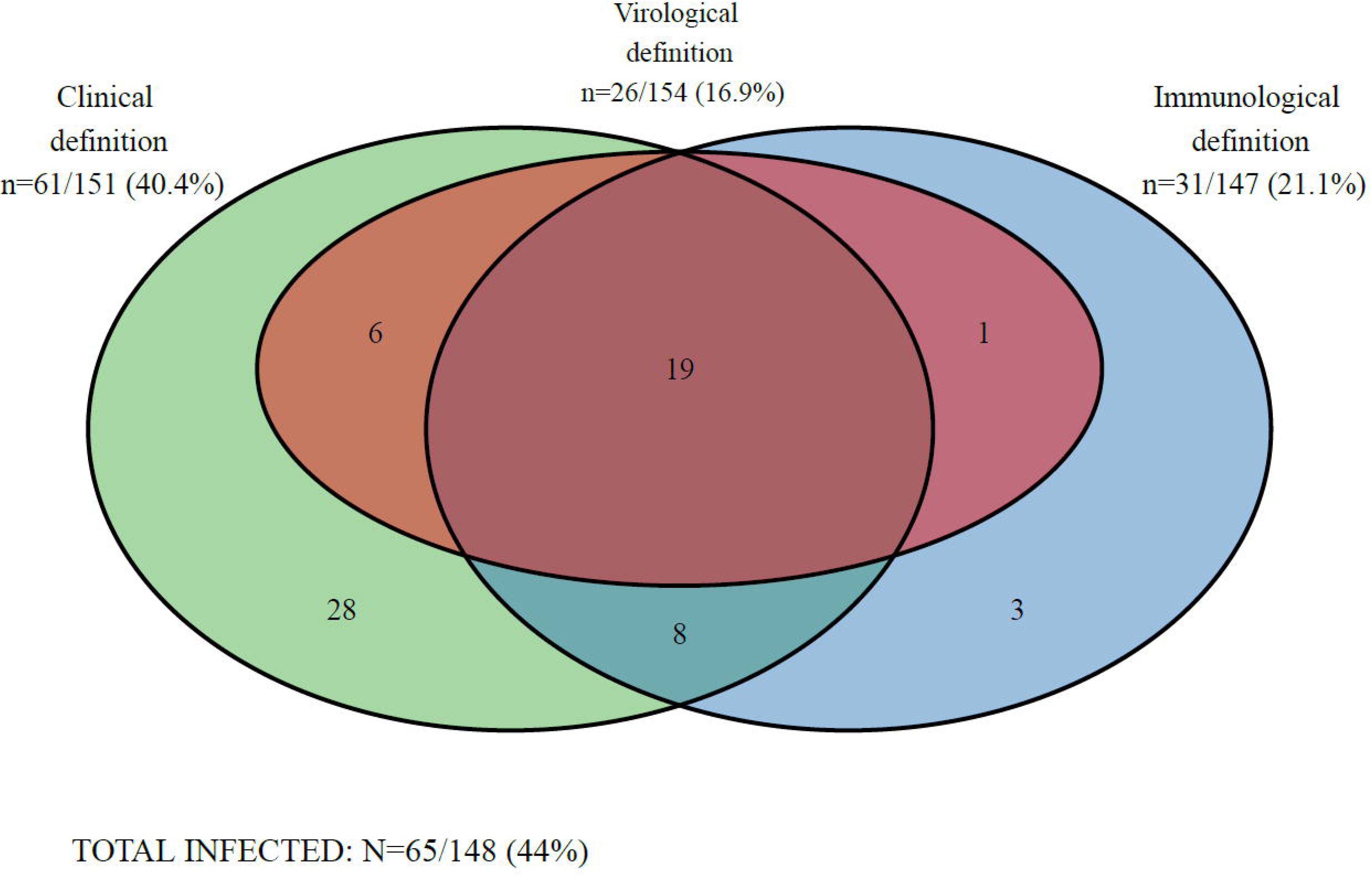
Venn diagram of the clinical, virological and immunological outcomes among the 154 contacts included in the CoV-CONTACT cohort. SARS-CoV-2 infection could be determined for 148/154 contacts (missing data for immunological and clinical outcomes (n=2), missing data for clinical outcome (n=1), missing data for immunological outcome (n=3).

**Figure 2.**
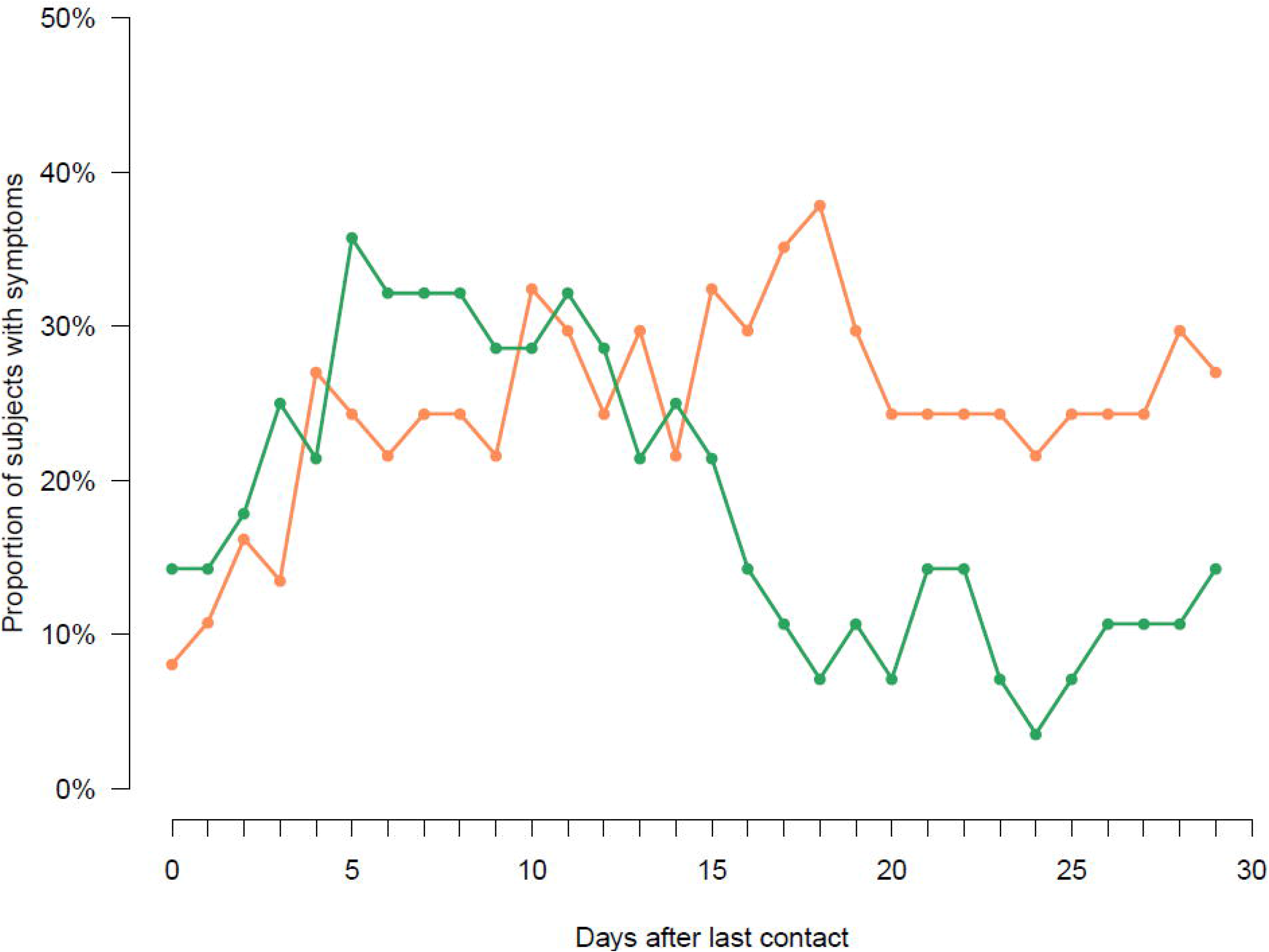
Proportions of symptomatic contact subjects among the 154 contacts included in the CoV-CONTACT cohort. The orange curve corresponds to contacts subjects with confirmed SARS-CoV-2 infection (/.e., virologically- or immunologically-proven, n=37). The green curve corresponds to contacts subjects with possible SARS-CoV-2 infection (/.e., clinically-suspected without viro-immunological confirmation, n=28).

### Proportion of contacts with SARS-CoV-2 infection

At D30, among the 148 contacts with available data, 65 met the criteria of confirmed or possible SARS-CoV-2 infection (43.9%, 95%CI [35.9%; 52.3%]), confirmed in 37 (25.0%, 95%CI [18.4%; 32.9%]), and possible (/.e., only clinically-suspected) in 28 (18.9%, 95%CI [13.2%; 26.5%]). Figure S1 presents the different clusters of exposure from the 44 index subjects. Among the 28 contacts with possible SARS-CoV-2 infection, multiplex RT-PCR for other respiratory viruses could be performed in 21 and was negative for 19 patients and positive for two (one bocavirus and one rhinovirus). During followup, one contact with confirmed SARS-CoV-2 infection was hospitalized. There was no hospitalization for SARS-CoV-2 infection reported in their household contacts.

### Factors associated with SARS-CoV-2 infection

In the multivariable analysis, the variables associated with SARS-CoV-2 infection were being a pharmacist or administrative assistant (OR=3.8, CI95%=1.3; 11.2, p=0.01) and having a contact with a SARS-CoV-2-infected patient (OR=2.6, CI95%=1.23; 5.9, p=0.02).

The results of the sensitivity analysis excluding contacts having only a possible SARS-CoV-2 infection provided similar results except for pharmacist or administrative assistants’ function (Table S2).

### Viral dynamics in infected contacts

The viral load as a function of time since symptom onset reached a maximum at 8.8 logio copies/ml 4 days after symptom onset followed by a decline afterwards (Figure 3). Of note, 7/25 subjects had a positive SARS-CoV-2 nasopharyngeal swab before the symptoms onset and the first positive nasopharyngeal swab was observed as early as six days before symptoms onset. In eight subjects, the positive swab was preceded by one negative swab and in two of them, the negative swab was done after the symptom onset.

**Figure 3.**
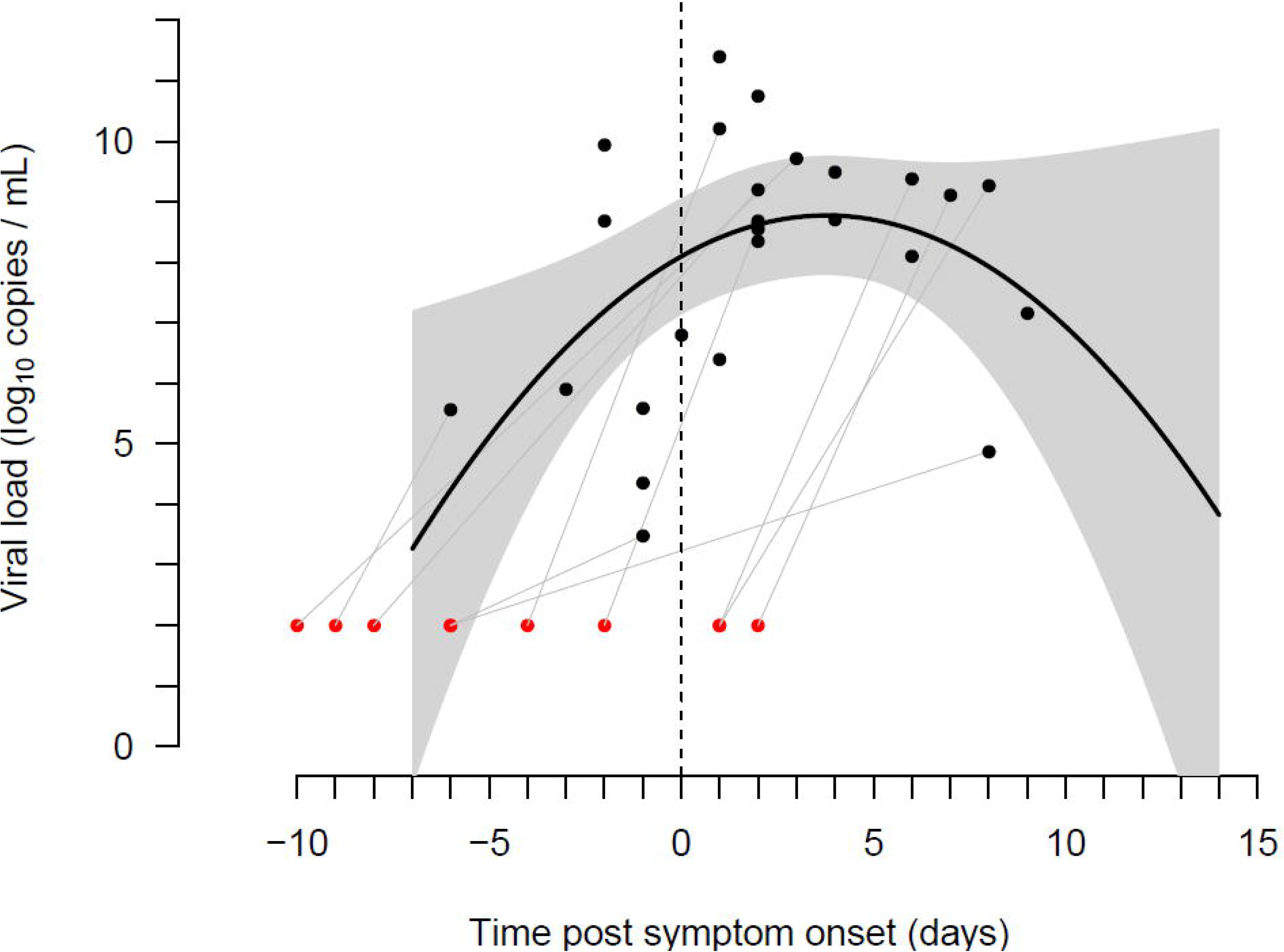
SARS-CoV-2 viral load in the first positive SARS-CoV-2 nasopharyngeal swab as a function of time since symptom onset, in the 25 healthcare workers with a positive RT-PCR and who met the definition of clinical infection. The first day when a specific symptom and a general symptom had been reported for two consecutive days was considered as the time of symptom onset. The black dots show the viral loads at the first positive RT-PCR for each subject. The red dots show the time of the previous negative RT-PCR when present (8 subjects). The black curve is the best fitting second-order polynomial for viral load as a function of time since symptom onset, and the shaded area is the 95% confidence interval.

## Discussion

In this prospective cohort of high-risk exposed HCWs, between 25% and 44% of subjects acquired SARS-CoV-2 infection at day 30, depending on the definition used to assess infection. Viral shedding occurred before symptoms onset in 27% of the SARS-CoV-2-positive subjects. The majority of HCWs were exposed to a SARS-CoV-2-infected colleague (62.5%), and a substantial proportion had a high-risk contact with a patient (46.1%). Exposure with a SARS-CoV-2-infected patient was significantly associated with SARS-CoV-2 infection (p=0.023).

The HCWs included in our analysis reflected the diversity of the hospital workers, [17] offering an ideal perspective to analyze the risks of infections encountered in a hospital. Following universal masking for HCWs on March, 18^th^ in our hospital, high-risk exposure to SARS-CoV-2-positive patients dropped from 68.2% to 11.8%, and high-risk exposure to SARS-CoV-2-positive colleagues became predominant, increasing from 31.8% to 88.2% and making colleagues-to-colleagues transmission a potentially major route of infection [7]. The profession of the contact subject was associated with infection, but we did not find any association with the type of activities of the HCWs. Although it is likely that activities involving a close contact with patients favor infection, such association might have been masked as HCWs could also be infected by colleagues.

One of the strengths of our study is its prospective design, with daily self-questionnaire to characterize symptoms onset and evolution, as well as repeated nasopharyngeal swabs to capture the time of infection after exposure, and serological assessments at inclusion and at day 30. The 10% seropositivity of HCWs at inclusion corresponds to the seroprevalence reported in the Paris area in the general population during this period, [18] and it is plausible that these subjects had been infected prior to the high-risk exposure identified in the study. Seroconversion was observed in about a third of exposed HCWs classified as infected, while their nasopharyngeal swabs were negative; most of these HCWs were symptomatic. Inversely a quarter of the HCWs with a SARS-CoV-2-positive nasopharyngeal swabs had no detectable antibodies at day 30, despite the use of two serological techniques. The prospective nature of our analysis allowed us to characterize the time to viral positivity in the 26 subjects with SARS-CoV-2-positive nasopharyngeal swabs and the relationship between the viral load and the time since symptom onset. Nasopharyngeal viral load could be positive before symptom onset, with the first positive viral load obtained as early as six days before symptom onset, consistent with the estimated 6-day incubation period previously reported [19, 20].

In addition to these confirmed infections, we also considered possible infections, as defined by the presence of one general and one specific symptoms for two consecutive days in subjects with neither a positive PCR nor seroconversion. Our definition is stricter than what was done in other studies [17, 21-23]. Interestingly we found that the kinetics of symptom onset was very similar in confirmed and possibly infected subjects, with 20-35% of subjects presenting symptom between day 4 and day 15 after their last high-risk exposure. The possibly infected patients had a lower prevalence of symptoms as time went by, with a rapid reduction of prevalence around day 15, while subjects with confirmed infection had a prevalence of symptoms that remained larger than 20% until day 30. The faster clearance of symptoms in possible infections suggests that these patients had a milder disease, which would be consistent with the fact that their nasopharyngeal swabs remained negative and that they did not exhibit detectable antibodies. Future analyses, that will include the probabilistic analysis of their serial negative nasopharyngeal swab results, as well as the study of the immune cellular response, will provide more conclusive evidence on their infection status [24].

A major limitation of our study is the absence of whole genome sequencing comparing the virus of the index subject and SARS-CoV-2-infected HCW. Therefore, the network of exposures and infection only suggests that the infection in a subject is the consequence of a high-risk exposure. However, sequencing would be restricted to RT-PCR positive subjects, which only represent 40% (26/65) of our population of confirmed and possible infections. Another limitation was that the type of contacts observed in the study has been modified by universal masking implemented on March, 18^th^, 2020. After this date, most at risk contacts were between two HCWs, which were less likely, but not unlikely, to result in SARS-CoV-2 transmission.

All together, the rate of transmission observed in HCWs after high-risk exposure, which could be as large as 44%, and close to a recent report [25], strengthens the conclusion that universal masking of HCW, both during contacts with patients and colleagues, and at all times, is essential to prevent HCWs infection and maintain hospital capacities during outbreaks [26].

## Data Availability

Data sharing will be considered upon request.

## Contributors

ST, CB, XD, YY, JCL, JB, PB and BL designed the experiments. ST, MT, LA, AL and XD included subjects. NH, CC, DD, SvW and BL performed the biological analyses. ST, CB, PM, FB, JG and XD analyzed the data. ST, CB, PM, FB, JG CC, DD, SvW, BL and XD interpreted the results. ST, CB, FB, and XD wrote the Article. All authors reviewed and approved the manuscript before submission.

## Funding

The study was founded by Reacting, the French Ministry of Health (PHRC-N COVID-19, project PHRC-20-0242) and the European Commission (RECoVer, grant agreement 101003589). Some authors received financial support of the national research agency (ANR) through the ANR-Flash calls for COVID-19 (TheraCoV ANR-20-COVI-0018)

## Declaration of interests

The authors have no commercial or other associations that might pose a conflict of interest.

## Acknowledgments

CoVCONTACT study group

**<u>Principal investigator:</u>** Duval Xavier

**<u>Steering Committee:</u>** Burdet Charles, Duval Xavier, Lina Bruno, Tubiana Sarah, Van Der Werf Sylvie

**<u>CoV-CONTACT Clinical Centers:</u>** Abad Fanny, Abry Dominique, Alavoine Loubna, Allain Jean-Sébastien, Amiel-Taieb Karline, Audoin Pierre, Augustin Shana, Ayala Sandrine, Bansard Hélène, Bertholon Fréderique,Boissel Nolwenn, Botelho-Nevers Elisabeth, Bouiller Kévin, Bourgeon Marilou, Boutrou Mathilde, Brick Lysiane, Bruneau Léa, Caumes Eric, Chabouis Agnès, Chan Thien Eric, Chirouze Catherine, Coignard Bruno, Costa Yolande, Costenoble Virginie, Cour Sylvie, Cracowski Claire, Cracowski Jean Luc, Deplanque Dominique, Dequand Stéphane, Desille-Dugast Mireille, Desmarets Maxime, Detoc Maelle, Dewitte Marie, Djossou Felix, Ecobichon Jean-Luc, Elrezzi Elise, Faurous William, Fortuna Viviane, Fouchard Julie, Gantier Emilie, Gautier Céline, Gerardin Patrick, Gerset Sandrine, Gilbert Marie, Gissot Valérie, Guillemin Francis, Hartard Cedric, Hazevis Beatrice, Hocquet Didier, Hodaj Enkelejda, Ilic-Habensus Emila, Jeudy A, Jeulin Helene, Kane Maty,Kasprzyk Emmanuelle, Kikoine John, Laine Fabrice, Laviolle Bruno, Lebeaux David, Leclercq Anne, Ledru Eric, Lefevre Benjamin, Legoas Carole, Legrand Amélie, Legrand Karine, Lehacaut Jonathan, Lehur Claire, Lemouche Dalila, Lepiller Quentin, Lepuil Sévérine, Letienne Estelle, Lucarelli Aude, Lucet Jean-Christophe, Madeline Isabelle, Maillot Adrien, Malapate Catherine, Malvy Denis, Mandic Milica, Marty-Quinternet Solène, Meghadecha Mohamed, Mergeay-Fabre Mayka,, Mespoulhe Pauline, Meunier Alexandre, Migaud Maria-Claire, Motiejunaite Justina, Nathalie Gay, Ngu,yen Duc, Oubbea Soumaya, Pagadoy Maëder, Paris Adeline, Paris Christophe, Payet Christine, Peiffer-Smadja Nathan, Perez Lucas, Perreau Pauline, Pierrez Nathalie, Pistone Thierry, Postolache Andreea, Rasoamanana Patrick, Reminiac Cécile, Rexah Jade, Roche-Gouanvic Elise, Rousseau Alexandra, Schoemaecker Betty, Simon Sandrine, Soler Catherine, Somers Stéphanie, Sow Khaly, Tardy Bernard, Terzian Zaven, Thy Michael, Tournier Anne, Tyrode Sandrine, Vauchy Charline, Verdon Renaud, Vernet Pauline, Vignali Valérie, Waucquier Nawal

**<u>Coordination and statistical analyses:</u>** Burdet Charles, Do Thi Thu Huong, Laouénan Cédric, Mentre France, Pauline Manchon, Tubiana Sarah, Dechanet Aline, Letrou Sophie, Quintin Caroline, Frezouls Wahiba

**<u>Virological lab:</u>** Le Hingrat Quentin, Houhou Nadhira, Damond Florence, Descamps Dianes, Charpentier Charlotte, Visseaux Benoit, Vabret Astrid, Lina Bruno, Bouscambert Maud, Van Der Werf Sylvie, Behillil Sylvie, Gaillanne Laurence, Benmalek Nabil, Attia Mikael, Barbet Marion, Demeret Caroline, Rose Thierry, Petres Stéphane, Escriou Nicolas, Barbet Marion, Petres Stéphane, Escriou Nicolas, Goyard Sophie

**<u>Biological center:</u>** Kafif Ouifiya, Piquard Valentine, Tubiana Sarah

**<u>Partners:</u>** RECOVER, REACTING, Santé Publique France (Coignard Bruno, Mailles Alexandra), Agences régionales de santé (Simondon Anne, Dreyere Marion, Morel Bruno, Vesval Thiphaine)

**<u>Sponsor:</u>** Inserm

Amat Karine, Ammour Douae, Aqourras Khadija, Couffin-Cadiergues Sandrine, Delmas Christelle,

Desan Vristi, Doute Jean Michel, Esperou Hélène, Hendou Samia, Kouakam Christelle, Le Meut

Guillaume, Lemestre Soizic, Leturque Nicolas, Marcoul Emmanuelle, Nguefang Solange, Roufai Layidé

**<u>Genetic:</u>** Laurent Abel, Sophie Caillat-Zucman

**<u>ClinicalTrial. Gov identification number:</u>** NCT04259892

## Supplementary Methods

### Data collection

In the case of exposure between colleagues working together, the beginning of the exposure period was fixed to 72 hours before the diagnosis of the SARS-CoV-2 infection of the index or the onset of the symptoms in the index, whichever occurred first.

### SARS-CoV-2 RT-PCR

Nasopharyngeal swabs were drawn by trained practitioners using Sigma Virocult® swabs (Medical Wire Instrument, UK) and processed within four hours after sampling. Nasopharyngeal swabs were manually discharged in conservation fluid according to the manufacturer recommendations. Viral RNA was extracted from 200 μL of discharge fluid with the MagNA Pure LC Total Nucleic Acid Isolation Kit – Large Volume (Roche Diagnostics) and eluted in 50 μL. Then, SARS-CoV-2 RT-PCR was performed using the RealStar® SARS-CoV-2 RT-PCR kit 1.0 (Altona Diagnostics) according to the manufacturer’s instructions. This assay allows the detection and differentiation of lineage B-betacoronavirus (B-BCoV), by targeting the E gene from B-BCoV, and SARS-CoV-2 specific RNA, by targeting the S gene. PCR assays were performed on an ABI 7500 plateform (Applied Biosystems®).

A signal with a cycle threshold value above 40 was considered as negative.

### Other respiratory pathogens RT-PCR

Detection of other respiratory pathogens was performed using the QIAstat-Dx Respiratory Panel (Qiagen), designed for the detection of adenovirus, bocavirus, coronavirus 229E (CoV 229E), CoV HKU1, CoVNL63, CoV OC43, human metapneumovirus A and B, influenza A (FLU A), FLU A H1, FLU A H3, FLU A H1N1/2009, influenza B, parainfluenza virus 1 (PIV 1), PIV2, PIV 3, PIV 4, human rhinovirus/enterovirus, respiratory syncytial virus A and B, *Bordetella pertussis, Chlamydophila pneumoniae*, and *Mycoplasma pneumoniae*. Approximately 300 μL of discharge fluid was tested according to the manufacturer’s instructions.

### LuLISA

The LuLISA N assay makes use of a nanobody fused to luciferase for the detection of IgG antibodies binding to the SARS-CoV-2 nucleoprotein (N). Sera were used at a 1:200 dilution. The luciferase signal was measured and expressed in Relative Light Units per second (RLU/s). The threshold of positivity (13,402 RLU/s) was based on the analysis of 231 pre-pandemic sera and defined as the mean + two standard deviations

## Supplementary Results

### Virological, immunological and clinical outcomes

The median number of nasopharyngeal swabs performed per contact was two (IQR 1;3), and the median time between the last high-risk exposure and the first SARS-CoV-2-positive nasopharyngeal swab was 6.5 days (IQR 4; 8).

Based on self-administered questionnaires, the most frequent symptoms being tiredness (74/151; 49.0%), headache (72/151; 47.7%) and myalgia (48/146; 32.9%) among general symptoms, and nasal congestion (52/148; 35.1%) and cough (51/149; 34.2%) among specific symptoms. The median duration of symptoms was 5.5 days (IQR 3; 9.2).

The median number of symptoms was 5 (IQR 4; 6) among the 28 contacts with possible SARS-CoV-2 infection and was 6 (IQR 4; 10) among those with confirmed SARS-CoV-2 infection.

### Factors associated with SARS-CoV-2 infection

Exposure to a SARS-CoV-2-infected patient was at higher risk of SARS-CoV-2 infection than exposure to a SARS-CoV-2 colleague (p=0.023, Table 1). Nurses, nurse assistants, physiotherapists and hospital students had a higher risk than medical staff (p=0.04), as did pharmacists and administrative assistants (p=0.033). In the multivariable analysis, the c-statistic of this final model was 0.67 (95%CI=0.58; 0.76) and the p-value of the Hosmer-Lemeshow test was 0.99, showing no model misspecification.

**Supplementary Figure S1.**
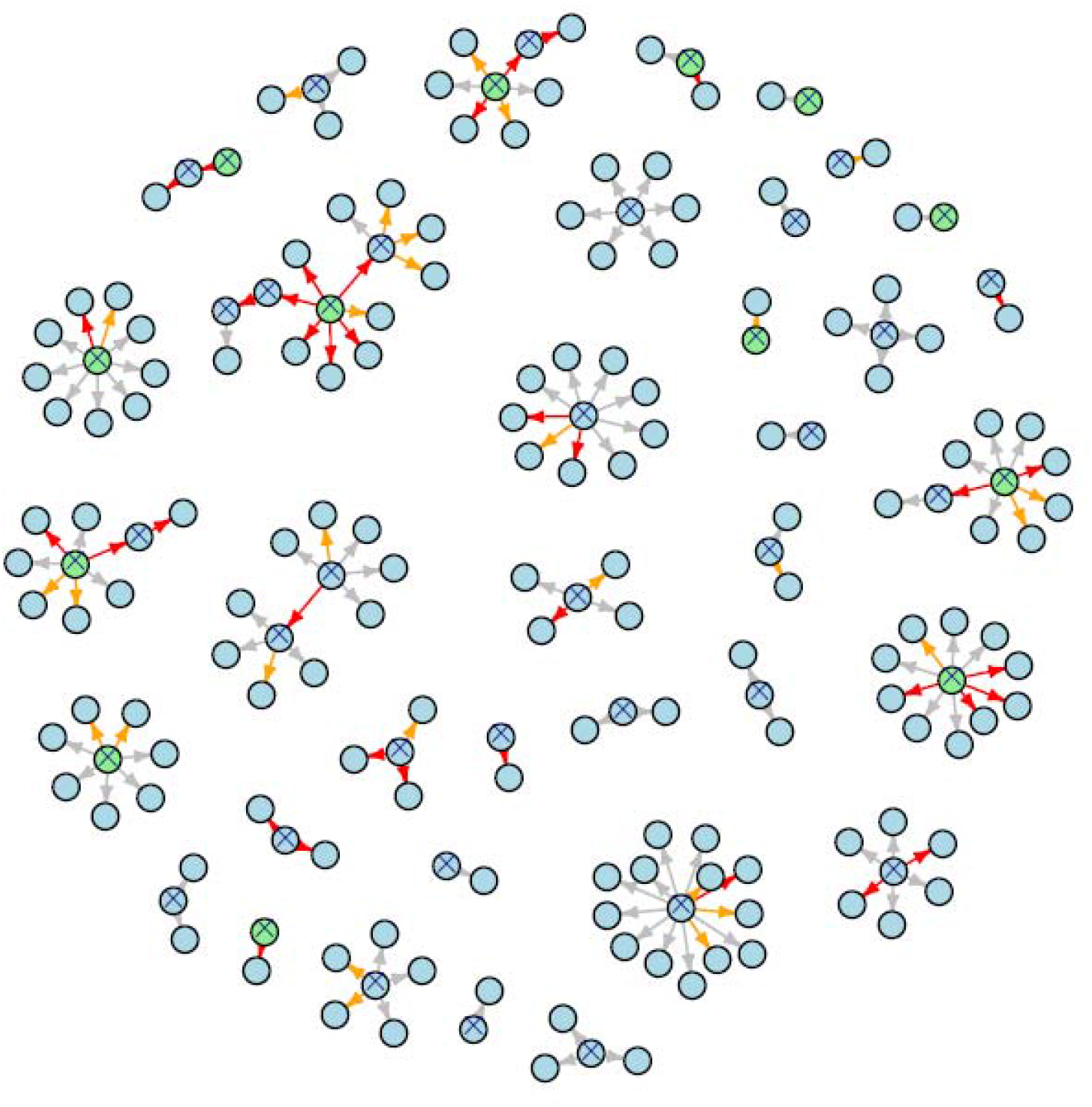
Structure of the contact network between the 44 index subjects and the 154 healthcare workers contact subjects with high-risk exposure to SARS-CoV-2 included in the CoV-CONTACT cohort.

Blue circles are health-care workers (either index or contact subjects), green circles are SARS-CoV-2 infected index patients. Circles with crosses represent the COVID-19 index subjects.

The arrows are the exposures; the red ones are the confirmed infections *(i.e*., virologically- or immunologically-proven, n= 37), the orange ones are the possible infections *(i.e*., clinically-suspected without viro-immunological confirmation, n=28).

**Supplementary Table S1.** Immunological results obtained by the two serological methods (Lulisa N and EuroIMMUN) in the 147 healthcare workers contact subjects with available data, following a high-risk exposure to SARS-CoV-2 included in the CoV-CONTACT cohort.

|  |  | Euroimmun technique |  |  |
| --- | --- | --- | --- | --- |
|  |  | No seroconversion | Seroconversion | Total |
| Lulisa N technique | No seroconversion | 116 | 2 | 118 |
|  | Seroconversion | 3 | 26 | 29 |
|  | Total | 119 | 28 | 147 |

**Supplementary Table S2.**
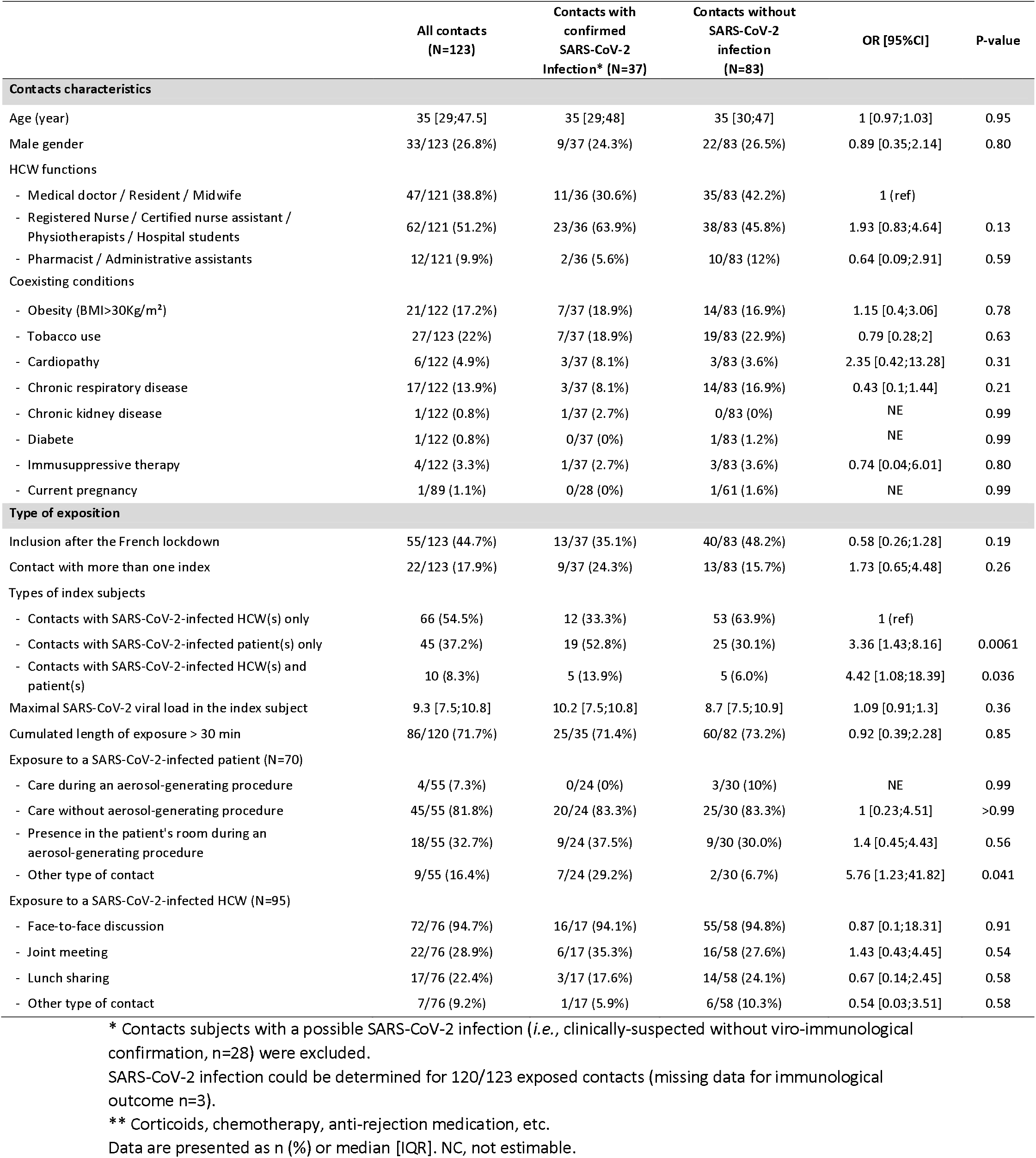
Characteristics of the 123 healthcare workers contact subjects with confirmed SARS-CoV-2 infection and without infection following a high-risk exposure included in the CoV-CONTACT cohort.

## Notes

### Competing Interest Statement

The authors have declared no competing interest.

### Clinical Trial

NCT04259892

### Author Declarations

The study was approved by the French National Data Protection Commission (approval #920102), and the French Ethics committee (CPP-Ile-de-France-6, #2020-A00280-39) and all subjects provided written informed consent.

